# Analysis of rare coding variants in 470,000 exome-sequenced subjects characterises contributions to risk of type 2 diabetes

**DOI:** 10.1101/2023.10.23.23297410

**Authors:** David Curtis

## Abstract

**Aims:** To follow up results from an earlier study using an extended sample of 470,000 exome-sequenced subjects to identify genes associated with type 2 diabetes (T2D) and to characterise the distribution of rare variants in these genes.

**Materials and methods:** Exome sequence data for 470,000 UK Biobank participants was analysed using a combined phenotype for T2D obtained from diagnostic and prescription data. Gene-wise weighted burden analysis of rare coding variants in the new cohort of 270,000 samples was carried out for the 32 genes previously significant with uncorrected p < 0.001 along with 7 other genes previously implicated in T2D. Follow-up studies of *GCK, GIGYF1, HNF1A* and *HNF4A* used the full sample of 470,000 to investigate the effects of different categories of variant.

**Results:** No novel genes were identified as exome wide significant. Rare loss of function (LOF) variants in *GCK* exerted a very large effect on T2D risk but more common (though still very rare) nonsynonymous variants classified as probably damaging by PolyPhen on average approximately doubled risk. Rare variants in the other three genes also had large effects on risk.

**Conclusions:** In spite of the very large sample size, no novel genes are implicated. Coding variants with an identifiable effect are collectively too rare be generally useful for guiding treatment choices for most patients. The finding that some nonsynonymous variants in *GCK* affect T2D risk is novel but not unexpected and does not have obvious practical implications.

This research has been conducted using the UK Biobank Resource.

## Introduction

A previous study carrying out gene-based weighed burden analysis of rare coding variants using 200,000 exome-sequenced UK Biobank participants identified three genes associated with type 2 diabetes (T2D) at exome-wide significance, *GCK, HNF4A* and *GIGYF1* (Curtis, 2021). While *GCK, HNF4A* were already well-recognised as causes of maturity onset diabetes of the young (MODY), the implication of *GIGYF1* was novel though was quickly confirmed in another study which had access to sequence data from 379,000 UK Biobank participants(Bishay and Greenfield, 2016; Naylor, Johnson and Gaudio, 2018; Deaton *et al*., 2021). Although these three were the only genes which achieved exome-wide significance, a total of 32 genes were significant with an uncorrected p value < 0.001 whereas, given that there were 20,384 informative genes, only 20 would be expected by chance. Additionally, a number of these genes appeared to be of potential interest from a biological point of view. Of note, a number of other genes with well-established roles in T2D failed to produce strong evidence of association using the weighted burden analysis, consisting of *HNF1A, HNF1B, ABCC8, INSR, MC4R, SLC30A8* and *PAM*.

Subsequently, rare variant analyses using multiple different phenotypes were carried out in larger numbers of exome sequenced participants from the same UK Biobank cohort and some of the phenotypes studied included T2D and related conditions (Backman *et al*., 2021; Wang *et al*., 2021).

Exome sequence data for a full set of 470,000 participants has now been made more widely available and the current study carried out weighted burden analysis in the new samples of the genes significant at p < 0.001 in the previous study, along with the other T2D implicated genes mentioned above. This study aimed to test for evidence of association and to compare results with those obtained from the multiple phenotype studies referred to above, as well as to characterise the effects of different categories of coding variant on risk in implicated genes.

## Materials and Methods

The methods used were essentially the same as those described previously and are briefly repeated here for the reader’s convenience.

UK Biobank participants are volunteers intended to be broadly representative of the UK population and are not selected on the basis of having any health condition. UK Biobank had obtained ethics approval from the North West Multi-centre Research Ethics Committee which covers the UK (approval number: 11/NW/0382) and had obtained informed consent from all participants. The UK Biobank approved an application for use of the data (ID 51119) and ethics approval for the analyses was obtained from the UCL Research Ethics Committee (11527/001). The UK Biobank Research Analysis Platform was used to access the Final Release Population level exome OQFE variants in PLINK format for 469,818 exomes which had been produced at the Regeneron Genetics Center using the protocols described here: https://dnanexus.gitbook.io/uk-biobank-rap/science-corner/whole-exome-sequencing-oqfe-protocol/protocol-for-processing-ukb-whole-exome-sequencing-data-sets (Backman *et al*., 2021). All variants were then annotated using the standard software packages VEP, PolyPhen and SIFT (Kumar, Henikoff and Ng, 2009; Adzhubei, Jordan and Sunyaev, 2013; McLaren *et al*., 2016). To obtain population principal components reflecting ancestry, version 2.0 of *plink* (https://www.cog-genomics.org/plink/2.0/) was run with the options -- *maf 0*.*1 --pca 20 approx* (Chang *et al*., 2015; Galinsky *et al*., 2016).

The T2D phenotype was defined in the same way as previously and was determined from three sources in the dataset: self-reported diabetes or type 2 diabetes (but not type 1 or gestational diabetes); reporting taking any of a list of named medications commonly used to treat T2D in the UK (https://www.diabetes.co.uk/Diabetes-drugs.html); having an ICD10 code for non-insulin-dependent diabetes mellitus in hospital records or as a cause of death (Curtis, 2021). Subjects in any of these categories were deemed to be cases while all other subjects were taken to be controls. In the primary analyses to implicate specific genes, attention was restricted to participants not included in the earlier study, consisting of 19,701 cases and 249,581 controls. For the subsequent analyses using the whole sample there were 33,629 cases and 436,136 controls.

The SCOREASSOC program was used to carry out a weighted burden analysis to test whether, in each gene, sequence variants which were rarer and/or predicted to have more severe functional effects occurred more commonly in cases than controls (Curtis, 2012, 2016, 2020). Attention was restricted to rare variants with minor allele frequency (MAF) <= 0.01 in cases or controls or both. As previously described, variants were weighted by overall MAF so that variants with MAF >= 0.01 were given a weight of 1 while very rare variants with MAF close to zero were given a weight of 10. Variants were also weighted according to their functional annotation using the GENEVARASSOC program, which was used to generate the input files for weighted burden analysis by SCOREASSOC. Variants predicted to cause complete loss of function (LOF) of the gene were assigned a weight of 100. Nonsynonymous variants were assigned a weight of 5 but if PolyPhen annotated them as possibly or probably damaging then 5 or 10 was added to this and if SIFT annotated them as deleterious then 20 was added. The full set of weights and categories is displayed in Table 1 of the previous study (Curtis, 2021). As described previously, the weight due to MAF and the weight due to functional annotation were multiplied together to provide an overall weight for each variant. Variants were excluded if there were more than 10% of genotypes missing in the controls and cases or if the heterozygote count was smaller than both homozygote counts in controls and cases. If a subject was not genotyped for a variant then they were assigned the subject-wise average score for that variant. For each subject a gene-wise weighted burden score was derived as the sum of the variant-wise weights, each multiplied by the number of alleles of the variant which the given subject possessed.

**Table 1.**
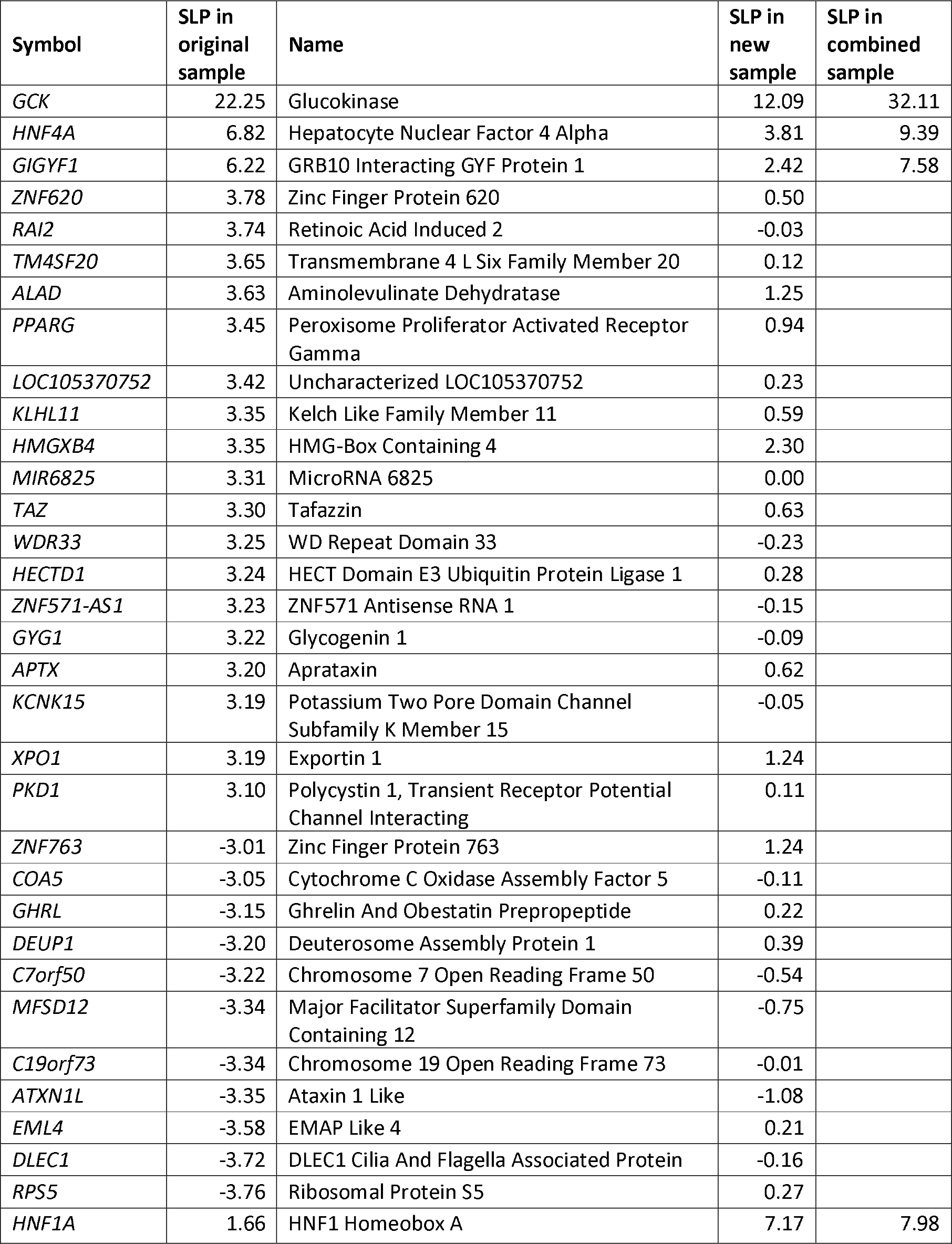
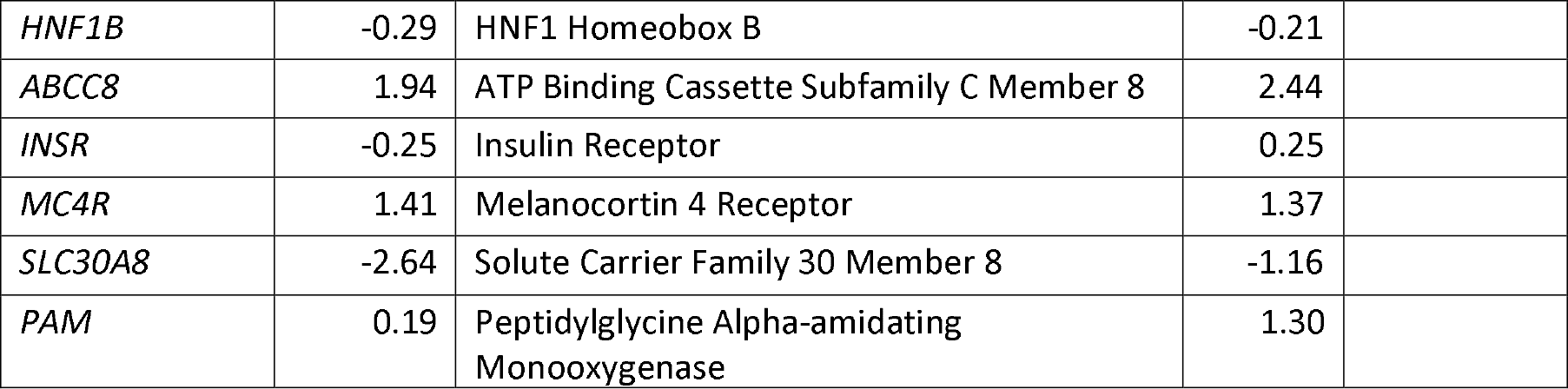
Results of gene-wise weighted burden analysis of rare variants in the original sample of 200,000 participants, the new sample of 270,000 and, for genes of interest, in the combined sample of 470,000.

Analyses were restricted to the 32 genes significant at p < 0.001 in the previous study along with the other 7 listed above as being previously implicated in T2D. For each gene, logistic regression analysis was carried out with T2D as the dependent variable including the first 20 population principal components and sex as covariates and a likelihood ratio test was performed comparing the likelihoods of the models with and without the gene-wise burden score. This is a test for association between the gene-wise burden score and caseness and the statistical significance was summarised as a signed log p value (SLP), which is the log base 10 of the p value given a positive sign if the score is higher in cases and negative if it is higher in controls. Since only 39 genes were analysed, after correcting for multiple testing a gene could be declared statistically significant if it achieved an SLP with absolute value greater than -log10(0.05/39) = 2.89 using the new samples.

Follow-up analyses were performed on all genes individually achieving this significance level and also *GIGYF1*. For this subset of genes the weighted burden analysis described above was carried out using the whole sample of 33,629 cases and 436,136 controls. Additionally, for each subject a count was obtained of the number of variants they carried falling into particular broad annotation categories, such as LOF, protein altering, etc. The full list of these categories is shown in Supplementary Table 1. These counts were entered into a multiple logistic regression analysis with T2D as the dependent variable and again including sex and 20 principal components as covariates in order to elucidate the contribution of different types of variant to the overall evidence for association. The odds ratios (ORs) associated with each category were estimated along with their standard errors and the Wald statistic was used to obtain a p value. This p value was converted to an SLP, again with the sign being positive if the OR was greater than 1, indicating that variants in that category tended to increase risk.

Data manipulation and statistical analyses were performed using GENEVARASSOC, SCOREASSOC and R (R Core Team, 2014; Curtis, 2016, 2020).

## Results

Table 1 shows the results of the primary analysis, presenting the SLPs obtained in the previous study along with those obtained in the new sample. Of the genes showing evidence for association in the previous study, only *GCK* (SLP = 12.09) and *HNF4A* (SLP = 3.81) are formally significant after correction for multiple testing, while *GIGYF1* yields SLP = 2.42 and none of the other genes previously with p < 0.001 shows evidence for association in the new sample. Of the 7 genes implicated in T2D in earlier studies, only *HNF1A* (SLP = 7.17) is formally statistically significant.

The four genes named above were carried forward for secondary analyses. The original study considered 20,384 genes, meaning that for a gene-wise result to be considered exome-wide significant the magnitude of the SLP obtained should exceed -log10(0.05/20384) = 5.61. For the four genes carried forward, the results of weighted burden analysis in the entire sample of 33,629 cases and 436,136 controls are also shown in Table 1 and it can be seen that all four of these genes produce results which would be regarded as exome-wide significant in the full sample.

In order to gain insights into the effects of different categories of variant within these four genes of interest, counts for variants of each category in each subject were entered into multiple logistic regression analysis along with sex and 20 principal components as covariates. These results are shown in Table 2 and are summarised briefly as follows.

**Table 2.**
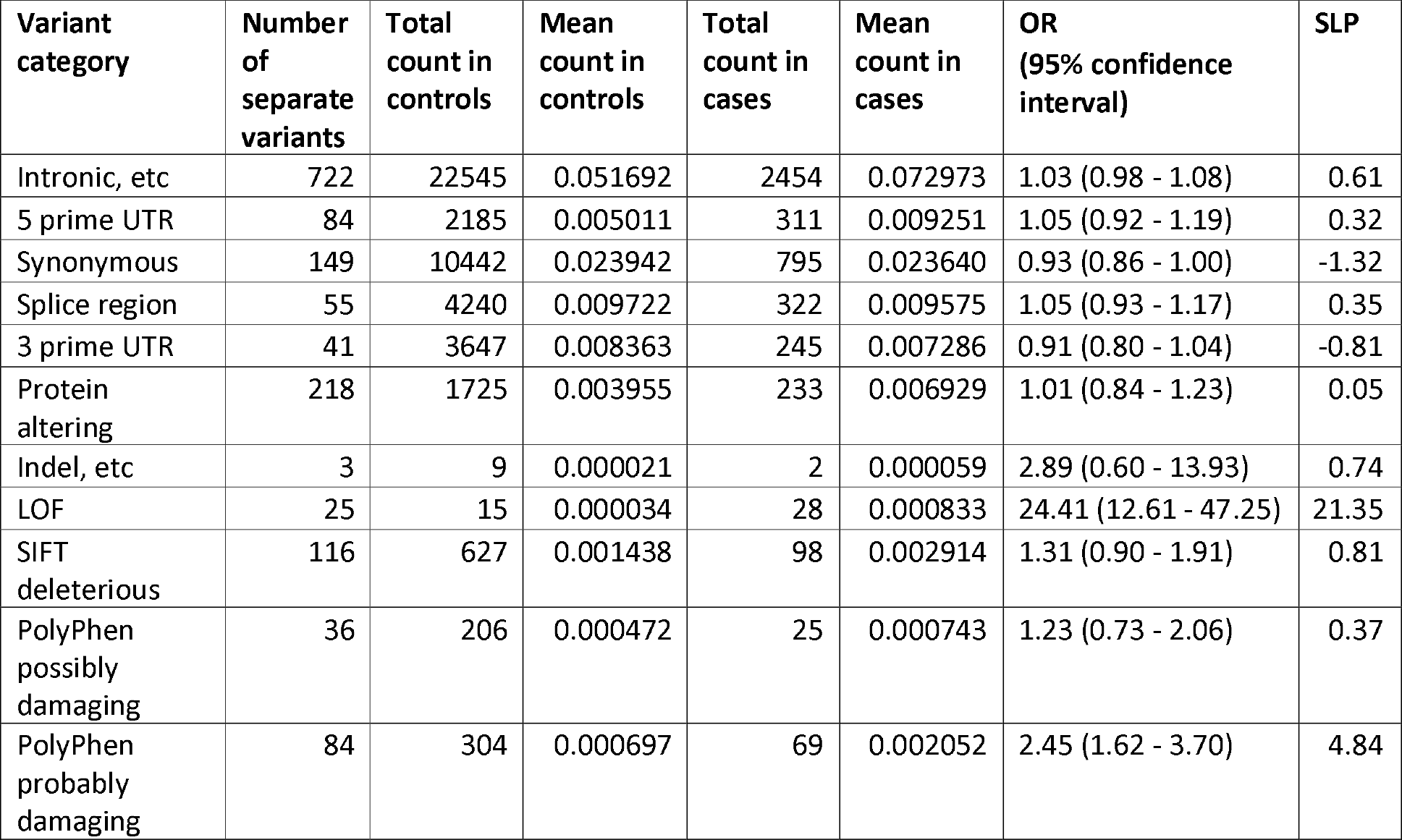
Results from logistic regression analysis including principal components and sex as covariates showing the contribution different categories of variant within each gene make to risk of hyperlipidaemia. Odds ratios for each category are estimated and the strength of evidence for an effect is expressed as the SLP. Table 2A Results for *GCK*.

| Variant category | Number of separate variants | Total count in controls | Mean count in controls | Total count in cases | Mean count in cases | OR (95% confidence interval) | SLP |
| --- | --- | --- | --- | --- | --- | --- | --- |
| Intronic, etc | 722 | 22545 | 0.051692 | 2454 | 0.072973 | 1.03 (0.98 - 1.08) | 0.61 |
| 5 prime UTR | 84 | 2185 | 0.005011 | 311 | 0.009251 | 1.05 (0.92 - 1.19) | 0.32 |
| Synonymous | 149 | 10442 | 0.023942 | 795 | 0.023640 | 0.93 (0.86 - 1.00) | -1.32 |
| Splice region | 55 | 4240 | 0.009722 | 322 | 0.009575 | 1.05 (0.93 - 1.17) | 0.35 |
| 3 prime UTR | 41 | 3647 | 0.008363 | 245 | 0.007286 | 0.91 (0.80 - 1.04) | -0.81 |
| Protein altering | 218 | 1725 | 0.003955 | 233 | 0.006929 | 1.01 (0.84 - 1.23) | 0.05 |
| Indel, etc | 3 | 9 | 0.000021 | 2 | 0.000059 | 2.89 (0.60 - 13.93) | 0.74 |
| LOF | 25 | 15 | 0.000034 | 28 | 0.000833 | 24.41 (12.61 - 47.25) | 21.35 |
| SIFT deleterious | 116 | 627 | 0.001438 | 98 | 0.002914 | 1.31 (0.90 - 1.91) | 0.81 |
| PolyPhen possibly damaging | 36 | 206 | 0.000472 | 25 | 0.000743 | 1.23 (0.73 - 2.06) | 0.37 |
| PolyPhen probably damaging | 84 | 304 | 0.000697 | 69 | 0.002052 | 2.45 (1.62 - 3.70) | 4.84 |

Table 2A shows that LOF variants in *GCK* exert a substantial risk of T2D, with OR over 20, but that nonsynonymous variants classified as probably damaging by PolyPhen also increase risk, with OR estimated as 2.45. Variants in this latter category are observed 363 times in the sample of 470,000 participants, so occur in less than 1 in 1,000 people, whereas the LOF variants are rarer still, being seen only 43 times.

Table 2B shows that LOF variants in *GIGYF1* are slightly commoner than in *GCK*, being seen 174 times, although they remain extremely rare. They have a more moderate effect on risk, with OR estimated as only 3.44. No other categories of variant have a clear effect on risk, though it is possible that variants classified has probably damaging by PolyPhen (SLP = 1.97) have a small effect (OR = 1.21).

**Table 2B.**
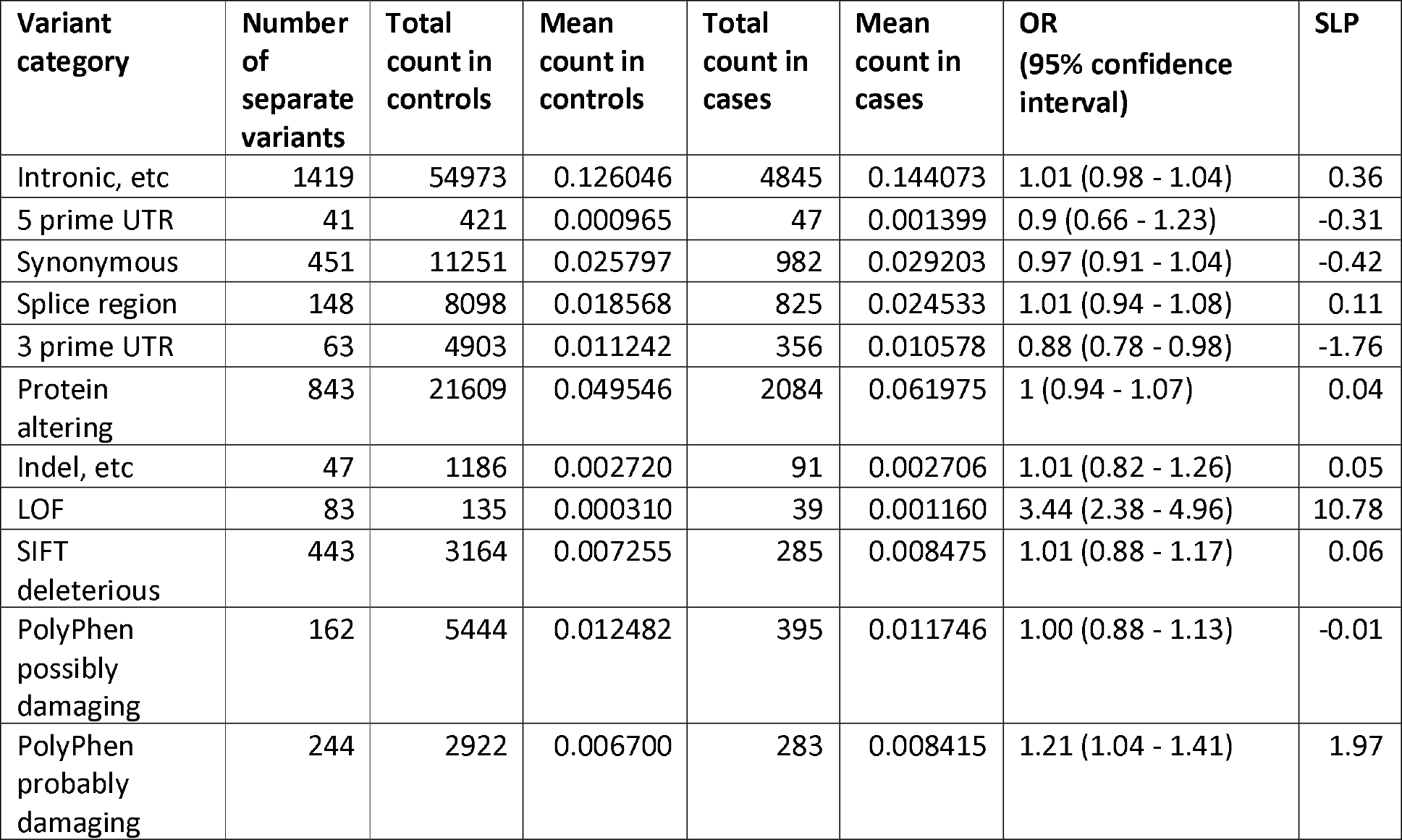
Results for *GIGFY1*.

Table 2C shows that LOF variants in *HNF1A* increase risk with OR = 4.88, but there may also be a modest effect of 5-prime UTR variants (SLP = 2.31, OR = 1.30) and/or variants classified as probably damaging by PolyPhen (SLP = 1.52, OR = 1.33).

**Table 2C.** Results for *HNF1A*.

| Variant category | Number of separate variants | Total count in controls | Mean count in controls | Total count in cases | Mean count in cases | OR (95% confidence interval) | SLP |
| --- | --- | --- | --- | --- | --- | --- | --- |
| Intronic, etc | 654 | 22154 | 0.050795 | 2087 | 0.062058 | 0.98 (0.93 - 1.03) | -0.47 |
| 5 prime UTR | 69 | 992 | 0.002274 | 136 | 0.004055 | 1.30 (1.08 - 1.56) | 2.31 |
| Synonymous | 223 | 3606 | 0.008268 | 369 | 0.010973 | 1.03 (0.92 - 1.15) | 0.22 |
| Splice region | 48 | 1304 | 0.002990 | 198 | 0.005888 | 1.09 (0.93 - 1.28) | 0.56 |
| 3 prime UTR | 48 | 775 | 0.001777 | 92 | 0.002736 | 1.06 (0.85 - 1.34) | 0.23 |
| Protein altering | 401 | 6623 | 0.015186 | 675 | 0.020072 | 1.09 (0.98 - 1.22) | 0.94 |
| Indel, etc | 6 | 19 | 0.000044 | 4 | 0.000119 | 3.12 (1.03 - 9.44) | 1.11 |
| LOF | 18 | 71 | 0.000163 | 24 | 0.000714 | 4.88 (3.01 - 7.88) | 10.36 |
| SIFT deleterious | 228 | 3004 | 0.006888 | 286 | 0.008505 | 0.95 (0.78 - 1.16) | -0.21 |
| PolyPhen possibly damaging | 96 | 1034 | 0.002371 | 98 | 0.002914 | 0.96 (0.75 - 1.23) | -0.12 |
| PolyPhen probably damaging | 126 | 1028 | 0.002357 | 114 | 0.003390 | 1.33 (1.02 - 1.72) | 1.52 |

Table 2D shows that LOF variants in *HNF4A* are extremely rare and do not have a detectable effect. Rather, the signal implicating this gene seems to come from variants classified as probably damaging by PolyPhen (SLP = 2.79, OR = 1.87) and two indel variants. These two variants consisted of 20:44428418GCCAACACAATGC>G (rs1349603952), observed in 4 controls and 2 cases, and 20:44424132A>AGCT (rs776489992), observed in 4 controls and 3 cases. Malacards lists 4 entries for rs776489992, with phenotypes MODY, MODY Type 1, T2D and Fanconi Renotubular Syndrome 4 with MODY (https://www.malacards.org/search/results?query=rs776489992). However there are no previous reports for rs1349603952.

**Table 2D.** Results for *HNF4A*.

| **Variant category** | **Number of separate variants** | **Total count in controls** | **Mean count in controls** | **Total count in cases** | **Mean count in cases** | **OR (95% confidence interval)** | **SLP** |
| --- | --- | --- | --- | --- | --- | --- | --- |
| Intronic, etc | 935 | 27523 | 0.063108 | 2774 | 0.082483 | 1 (0.96 - 1.04) | 0.01 |
| 5 prime UTR | 62 | 795 | 0.001823 | 57 | 0.001682 | 0.81 (0.61 - 1.07) | -0.87 |
| Synonymous | 192 | 11920 | 0.027332 | 1039 | 0.030897 | 1.02 (0.96 - 1.09) | 0.34 |
| Splice region | 51 | 2656 | 0.006090 | 319 | 0.009488 | 1.05 (0.93 - 1.20) | 0.38 |
| 3 prime UTR | 54 | 1089 | 0.002497 | 119 | 0.003541 | 0.9 (0.73 - 1.09) | -0.57 |
| Protein altering | 301 | 3225 | 0.007395 | 399 | 0.011865 | 1.08 (0.93 - 1.25) | 0.49 |
| Indel, etc | 2 | 8 | 0.000018 | 5 | 0.000149 | 9.76 (3.09 - 30.86) | 2.83 |
| LOF | 9 | 9 | 0.000021 | 1 | 0.000030 | 1.78 (0.27 - 11.70) | 0.28 |
| SIFT deleterious | 126 | 1400 | 0.003210 | 173 | 0.005144 | 1.34 (0.96 - 1.86) | 1.09 |
| PolyPhen possibly damaging | 40 | 738 | 0.001692 | 68 | 0.002022 | 0.88 (0.59 - 1.29) | -0.3 |
| PolyPhen probably damaging | 59 | 326 | 0.000747 | 69 | 0.002052 | 1.87 (1.26 - 2.78) | 2.79 |

## Discussion

These analyses provide very strong support for *GCK* as a risk gene for T2D while three other previously identified genes also achieve conventional levels of significance: *GIGYF1, HNF1A* and *HNF4A*. However, no novel genes are implicated. As mentioned previously, this dataset has been used for analyses of multiple phenotypes including some relating to T2D. which we can refer to as the Regeneron and AstraZeneca studies (Backman *et al*., 2021; Wang *et al*., 2021). The Regeneron study carried out a variety of single variant and gene-wise burden tests on 3,994 health-related traits to produce a total of about 2.3 billion tests, yielding a critical p value of 2.18e-11 (corresponding to SLP = 10.66), and reported 8,865 significant associations which are presented in their Supplementary Data 2 (Backman *et al*., 2021). 64 associations were reported between *GCK* and diabetes or related phenotypes, with the most significant being with glycated haemoglobin HbA1c at p = 4.98e-22, equivalent to SLP = 21.30, whereas in the current study *GCK* yields SLP = 32.11. *GIGYF1* was associated with T2D at SLP = 12.34 and *HNF1A* with T2D at SLP = 12.58. However no association with a diabetes-related phenotype was reported for *HNF4A*, although it was associated with levels of sex hormone binding globulin (SHBG) at SLP = 41.85. For the AstraZeneca study, all gene-wise and variant-wise associations with 17,361 binary and 1,419 quantitative phenotypes are reported on the AstraZeneca PheWAS Portal at *https://azphewas.com* (Wang *et al*., 2021). This was accessed to find the most significant p value for any analysis of each of these genes with the phenotype “Union#E11#Type 2 diabetes mellitus” and Table 3 shows the results obtained compared with those for the current study. It can be seen that the current study again produces stronger evidence for association with *GCK*, with SLP = 32.11 versus SLP = 23.10 for the AstraZeneca study, whereas for the other three genes the strength of evidence for association is fairly similar between the two studies.

**Table 3.** Comparison of results from current study to those reported for the AstraZeneca study. The results for the AstraZeneca study are displayed as the equivalent SLP for the most significant result reported for that gene with the phenotype “Union#E11#Type 2 diabetes mellitus”.

| Gene | SLP for combined sample in current study | SLP for AstraZeneca study |
| --- | --- | --- |
| *GCK* | 32.11 | 23.10 |
| *GIGYF1* | 7.58 | 8.77 |
| *HNF1A* | 7.98 | 6.85 |
| *HNF4A* | 9.39 | 9.37 |

The fact that current study finds stronger evidence for association of *GCK* relative to the other analyses may reflect the fact that, for this gene, the pattern of effects due to different variant types does resemble the model which is assumed for the weighted burden analysis, with strong effects due to LOF variants and more moderate effects due to some nonsynonymous variants. However for the other three genes this pattern is not seen and hence for them the weighted burden analysis does not have advantages over more conventional variant pooling analyses.

It is also of interest to note that the evidence in favour of the association with T2D risk is considerably higher for *GCK* than for the other genes, and likewise the effect size of implicated variants is larger. It is tempting to speculate that this relates to the molecular mechanisms underlying the observed association. The product of *GCK*, glucokinase, is a low-affinity hexose kinase which acts as the rate limiting enzyme for glycolysis in pancreatic islet cells, as well as in some hepatocytes and neurons, meaning that it can be used by these cells as an indicator of blood glucose levels (Ogunnowo-Bada *et al*., 2014; McCrimmon, 2022). Thus, impaired functioning of glucokinase is expected to lead to reduced sensitivity to higher glucose levels and hence inadequate glycaemic control. By contrast, *GIGYF1, HNF1A* and *HNF4A* are involved with lower level cellular processes which have a less immediate impact in terms of producing diabetes as a phenotype. The product of *GIGFY1* binds to Grb10, a protein which regulates the response to insulin-like growth factor receptor signalling and it is associated with a number of different phenotypes in addition to T2D, including lipid-related phenotypes, education score, cognitive function and cystatin C levels (Giovannone *et al*., 2003; Backman *et al*., 2021; Jurgens *et al*., 2022; Chen *et al*., 2023). The products of *HNF1A* and *HNF4A* are transcription factors affecting the expression of large numbers of other genes and influencing development of the liver and pancreas (Xue *et al*., 2023). Biallelic variants in *HNF1A* can cause hepatocellular adenomas, while variants in *HNF4A* can cause Fanconi renotubular syndrome and are associated with SHBG levels (Bioulac-Sage, Sempoux and Balabaud, 2017; Backman *et al*., 2021; Lemaire, 2021). The fact that *GCK* has such a direct effect on contributing to the control of glucose levels may explain in part why LOF variants in it have a larger effect on the T2D phenotype than for other genes.

It could be argued that the work presented here highlights some of the limitations as well as strengths of analysing rare coding variants identified in exome-sequencing studies of large population cohorts. Because of the high prevalence of T2D, many thousands of cases are available for study but, as the results show, only a small fraction of these cases carry a variant in a category which can be identified as impacting risk. A number of genes which had previously been implicated in targeted studies do not in the current study yield evidence at conventional levels of statistical significance after correction for multiple testing. Although T2D has a high prevalence, many other clinically important phenotypes have a substantial genetic contribution to risk but with a lower prevalence and there would be insufficient case numbers present in an unselected cohort for similar approaches to be likely to yield any convincing novel rare variant associations. In order to identify genes involved in such conditions it would be necessary to carry out studies involving specifically recruited cases, perhaps also focusing on those from densely affected families where large-effect variants may be active. To support such initiatives, it would be helpful to strengthen methods to incorporate existing samples as controls rather than requiring that a matching set of controls be recruited and sequenced for each new set of cases. Using existing samples as controls has been helpful in other sequencing studies but requires careful alignment of methodologies to minimise artefacts (Singh and The Schizophrenia Exome Meta-Analysis (SCHEMA) Consortium, 2022).

If adequately sized samples are used, exome sequencing studies can identify genes in which damaging variants have large effects on risk of particular phenotypes. The main value of such studies is to implicate specific genes, and hence their protein products, as impacting the phenotype. This may ultimately lead to a better understanding of the molecular pathways involved in pathogenesis. However, because for non-Mendelian diseases identifiable variants are only seen in a very small proportion of cases, typically fewer than 1%, such approaches seem unlikely to be helpful to guide individual treatment interventions in most situations. The vast majority of patients would not carry a variant which could be clearly identified as causal, and even if such a variant were encountered this might not automatically have clear implications for treatment choices.

Exome-sequencing studies to date, including the current one, now fairly consistently show that the category of variant having the highest identifiable impact on phenotype consists of those variants which are predicted to cause loss of function of the gene, or haploinsufficiency. This is not to say that individual variants in other categories might not have larger effects, and of course the literature is replete with examples of these. However, in a situation where individual variants are extremely rare, as is expected for those with large effect, it becomes necessary to pool variants together in some form of burden analysis and currently available methods for prediction of impact of non-LOF variants on the function of the gene and/or protein product are not able to reliably discriminate those which are pathogenic from those without major effect. If for a given gene-phenotype pair we can discover that that LOF variants have a particular effect on increasing or decreasing risk then this may provide an important endpoint in terms of improved insight into the molecular pathways involved in pathogenesis. For example, this might be sufficient to flag up the protein as a possible drug target. However it is possible to argue that additional useful information could be gained from more intensive investigations to elucidate the effects of other types of variant. For example, if one can find that non-synonymous variants affecting particular protein domains tend to show evidence of association this might yield a more sophisticated understanding of disease mechanisms which again might potentially be exploited therapeutically.

The present study confirms the role of four previously implicated genes in risk of T2D. It also demonstrates that nonsynonymous variants in *GCK* which PolyPhen annotates as probably damaging on average approximately double risk of T2D, although as these variants are still very rare this finding may not have much in the way of practical applications. The results show the distributions of different categories of variant in these genes in the general population. Overall, the study provides some insights into what can be achieved from the analysis of exome sequence data and into some limitations of such approaches.

## Author contributions

DC conceived the project, carried out the analyses and wrote the manuscript.

## Data availability

The raw data is available on application to UK Biobank. Detailed results with variant counts cannot be made available because they might be used for subject identification. Scripts and relevant derived variables will be deposited in UK Biobank. Software and scripts used to carry out the analyses are also available at https://github.com/davenomiddlenamecurtis.

## Acknowledgments

This research has been conducted using the UK Biobank Resource. The author wishes to acknowledge the staff supporting the High Performance Computing Cluster, Computer Science Department, University College London. The author wishes to thank the participants who volunteered for the UK Biobank project.

---

**Supplementary Table 1.** The table shows the broad categories used for variant category specific analyses along with the annotations produced by VEP which were grouped into each category.

| Category | VEP annotation |
| --- | --- |
| Intronic etc. | feature_truncation, regulatory_region_variant, feature_elongation, regulatory_region_amplification, regulatory_region_ablation, TF_binding_site_variant, TFBS_amplification, TFBS_ablation, downstream_gene_variant, upstream_gene_variant, non_coding_transcript_variant, NMD_transcript_variant, intron_variant, non_coding_transcript_exon_variant |
| Five prime UTR | 5_prime_UTR_variant |
| Synonymous | synonymous_variant |
| Splice region | splice_region_variant |
| Three prime UTR | 3_prime_UTR_variant |
| Protein altering | protein_altering_variant, missense_variant |
| Indel etc. | inframe_deletion, inframe_insertion, transcript_amplification |
| LOF | frameshift_variant, stop_gained, transcript_ablation, splice_donor_variant, splice_acceptor_variant |
| SIFT deleterious | deleterious |
| PolyPhen possibly damaging | possibly_damaging |
| PolyPhen probably damaging | probably_damaging |

